# Keeping on target during a low-calorie diet: The experiences of participants in a type 2 diabetes reversal pilot

**DOI:** 10.1101/2022.08.02.22278322

**Authors:** Latoya Bartholomew, Nigel Unwin, Cornelia Guell, Madhuvanti M. Murphy

## Abstract

Rapid weight loss using a very low-calorie diet (VLCD) for 2 to 3 months followed by weight maintenance has been shown to lead to the restoration of normal glucose and insulin metabolism in people recently diagnosed with type 2 diabetes. We explored the barriers and facilitators of adhering to a VLCD for remission in an Afro-Caribbean population. Twenty-five participants completed an eight-week VLCD followed by a six-month structured weight maintenance phase. Semi-structured interviews and focus groups were thematically analysed. Cravings for ‘usual foods’ consumed by friends and family, stigma of diabetes and resulting lack of disclosure and the participants’ busy schedules were noted as predominant challenges throughout the study. In turn, social support and the participants’ internal drive were considered as key facilitators for success. VLCD is a feasible approach for type 2 diabetes remission. Self-motivation and supportive environments are however crucial in meeting and maintaining the weight loss goals.

## Introduction

Diabetes is a major global public health concern, with prevalence and mortality rates rising in both developed and developing countries alike [1, 2]. With a global prevalence estimated at 463 million adults as of 2019, these figures are projected to rise to 578 million by 2030, with type 2 accounting for 90% of all diagnosed cases [3]. Over the past decades the prevalence has risen faster in low and middle income countries when compared to high income countries [4, 5]. In developing countries in particular, adult diabetes numbers are likely to increase by 69% from 2010 to 2030 compared to 20% for developed countries [1].

The high burden of type 2 diabetes has been linked to several wide-ranging complications which affect all aspects of an individual’s quality of life. For the period 2000-2016, diabetes ranked among the three leading causes of death in the Region of the Americas [6], with the highest rates recorded in most countries of the English-speaking Caribbean [7]. In Barbados, approximately 19% of adults have the illness with the prevalence being the highest for those 65 years and older [8]. In the 2019 Global Burden of Disease Study, diabetes was ranked the third leading cause of death and the number one health problem to cause the most disability for Barbadians [9].

Parallel to the increasing rates of diabetes is the epidemic of obesity. It is well established that overweight/obesity is the major modifiable risk factor for type 2 diabetes. In a 10-year follow-up study, Field et al. [10] found that those with a body mass index (BMI) of 35.0 or more were 20 times more likely to develop diabetes than their same-sex peers with a BMI between 18.5 and 24.9. At the crux of these epidemics lie causes such as profound changes to work, transport and leisure, and changing physical and social environments including urbanisation leading to population changes in physical activity and poor nutrition [11]. Despite extensive health promotion initiatives and behaviour modification programmes, type 2 diabetes continues to be one of the most challenging chronic illnesses to manage [12], and requires lifestyle changes and adherence to complex daily regimes including dietary modification and physical activity [13-16].

Although sustained behaviour modification is an important part of the prevention and treatment of diabetes, it remains elusive for many individuals [16]. This is so as one’s dietary change can be affected by barriers related to self-discipline, emotions, knowledge, and access to social and financial resources. Moreover, the challenge of sustained behaviour modification is hinged on the influence of social factors on food accessibility and affordability, planning and preparation, and the central symbolic and emotional role food plays in one’s social life. Interventions that aim to improve metabolic control must be able to address such complexities.

Weight loss has long been recognized as a key component in producing optimal outcomes for patients with diabetes. Recent studies provide compelling evidence that persons with type 2 diabetes can return to normal glucose control through substantial weight loss achieved by adhering to a very low-calorie diet [17, 18]. Previous qualitative research however suggests that the adherence to such a diet is impacted by several barriers inclusive of tempting situations and environments, emotional distress, and high costs of healthy foods [19, 20]. This study thus explored the barriers and facilitators of adhering to an eight-week very low-calorie diet and maintaining the weight loss during the following six months.

## Materials and Methods

### Study Design

This work is part of a larger study that examined the feasibility of a very low-calorie diet and structured long-term support for inducing the remission of type 2 diabetes in Barbados. Following similar research conducted in the UK by Lim et al. [21] and Steven et al.[22], this study spanned an eight-month period, with the first two months being for the very low-calorie liquid diet, followed by six months of structured weight maintenance. Ethical approval was obtained from the University of the West Indies Institutional Review Board and the Ministry of Health of the Government of Barbados. Written informed consent was secured from all participants.

Through purposive sampling, a total of 25 Barbadians who were diagnosed with type 2 diabetes for six years or less, between the ages of 20-69 with a body mass index of ≥ 27 kg/m^2^ participated in the study. The 25 participants were divided into two cohorts: the first comprising of 13 participants and the second, 12 participants. This division of the participants aided the researchers in ensuring that the data collection process for the weight loss phase (i.e., the first eight weeks) was managed effectively. The first cohort began their low-calorie liquid diet in March 2015 and continued until May at which point the second cohort began their eight-week low-calorie liquid diet. All participants stopped their use of hypoglycaemic medication on day one of the low-calorie liquid diet. The liquid diet comprised of four shakes per day, each containing 190 calories, 23 grams carbohydrate, 10 grams protein and 7 grams fat. These shakes were supplied at no cost to the participants. Participants were also encouraged to consume three litres of water and to supplement their liquid diets with 250g of high fibre low carbohydrate vegetables daily.

Employing a ‘before and after’ design, the feasibility study assessed the primary outcome measures of a reduction in weight, fasting glucose, and estimated change in beta cell function. The secondary outcome measures such as reductions in the participants’ waist circumference, blood pressure, adverse lipid profile and use of diabetes and blood pressure medication were also assessed.

### Setting

Barbados is one of the most developed Caribbean economies that continues to make positive strides towards achieving its economic and Millennium Development Goals. Comprehensive health care is available to all citizens through the island’s two-tiered healthcare system comprising of publicly funded and private clinics. In the public sector, primary, secondary, and tertiary levels of healthcare are regulated by the Ministry of Health. Citizens with diabetes can access primary care through the Ministry’s nine polyclinics free of charge, or for a fee they can visit their private general practitioners [23, 24]. Through the Barbados Drug Benefit Service, citizens can access at any one time, a maximum of one month’s supply of free medication [24]. For those patients with uncontrolled diabetes or who are at risk of complications, they are referred to the Barbados Diabetes Foundation’s specialty clinic where they are managed at a fee for a six-month period after which time they are once again referred to the primary healthcare system. Although citizens have universal access to healthcare, the public system is fraught with challenges; these include long waiting times to be seen in the polyclinics, long waiting lists at the Queen Elizabeth Hospital, the main provider of acute secondary and tertiary care and medication supply problems in the polyclinics [24, 25].

### Data Collection and Analysis

Individual in-depth semi-structured interviews were conducted with the 25 participants during the three main data collection time-points: baseline, eight weeks, and eight months. Furthermore, two focus group sessions each comprising of six participants were conducted at the end of the eight-week low-calorie diet. Key ideas, issues and concerns the participants had having completed the requirements of this critical juncture of the intervention were further explored.

The individual interviews explored barriers and facilitators participants experienced during the intervention and ranged from 5-50 minutes in length, while the focus groups lasted no longer than an hour and thirty minutes. All but three interviews were recorded with the participants’ consent and transcribed verbatim by LB. Detailed notes were however taken in the instance where the participant wished not to be recorded. Confidentiality and anonymity were maintained by assigning pseudonyms (used below) and removing any information which could be used to identify the participants.

All transcripts were reviewed in their entirety and notes were taken, as the researchers familiarized themselves with the data set. They were checked for accuracy and imported into ATLAS.ti 8.0 data analysis software (ATLAS.ti Scientific Software Development GmbH, Berlin Germany). The data were subsequently analysed through thematic analysis. Utilizing an inductive approach, line by line coding was completed and initial individual codes were developed. Codes were then grouped, refined, and used to examine patterns, ambiguities, and outliers among the transcripts. A synopsis of the key themes, elucidated by relevant quotes are presented and discussed below.

## Results

Of the 25 respondents who participated in this study, 10 were men and 15 were women with a mean duration of diagnosed diabetes of 3.0 (2.1) years. Table 1 provides an overview of selected baseline demographic characteristics

**Table 1.** Selected baseline characteristics of participants (N=25)

| *Characteristic* | *Men (N=10)* | *Women (N=15)* | *All (N=25)* |
| --- | --- | --- | --- |
| **Age, years** | 51 (12) | 46 (9) | 48 (10) |
| **Ethnicity, %** |  |  |  |
| Black | 80 | 93 | 88 |
| White | 0 | 7 | 4 |
| Mixed | 20 | 0 | 8 |
| **Diabetes Treatment, %** |  |  |  |
| Diet only | 0 | 20 | 12 |
| Oral medication | 100 | 80 | 88 |
| **Time since diagnosis, years** | 3.0 (1.8) | 3.1(2.3) | 3.0 (2.1) |
| **Weight, kg** | 107.7 (22.3) | 92.3(17.6) | 98.5(20.7) |
| **Height, cm** | 177.7 (8.9) | 164.0(5.6) | 169.5 (9.7) |
| **BMS, kg/m<sup>2</sup>** | 33.9 (5.5) | 34.4 (6.6) | 34.2 (6.2) |

*Note.* Data given as mean (SD) unless specified otherwise

The thematic analysis of the individual and group interviews revealed distinct challenges and facilitators of adhering to a low-calorie diet and maintaining weight loss for type 2 diabetes remission. Though there was some variation in the barriers and facilitators across the study’s lifespan, five common themes were noted: (1) Barriers to lifestyle changes; (2) Situational Barriers (3) Attitudes towards diabetes; (4) Social Support and (5) Internal drive and self-motivation. These themes have been structured and discussed below under the three critical categories (i.e., baseline, eight weeks, six-month weight maintenance) of the study.

### Barriers to lifestyle changes

During the baseline interviews, participants noted the difficulty they experienced maintaining a healthy lifestyle. This difficulty, they explained, stemmed mainly from their inability to maintain a healthy diet due to busy schedules, family responsibilities, desk-bound sedentary jobs and frequent choice of convenience food and eating out.

Within their households, participants noted consistent struggles with temptations as their family members consumed foods that were not ‘diabetes friendly’. The female participants expressed the heightened challenges they experienced as they were the ones tasked with the responsibility of preparing meals. To address this challenge, they attempted to cook separate meals for themselves and their family, a task which they agreed was difficult to maintain in the long run because of their busy schedules. As such, they reverted to consuming the same meals (which were typically high in carbohydrates, sugars, and fats) prepared for other members of the household, which made them question their ability to stick to the very low-calorie liquid diet. They perceived that they would experience ‘withdrawal symptoms’, such as hunger pangs, having low energy, and experiencing excessive headaches during the eight-week diet since they could no longer consume their favourite foods and drinks.

Throughout the formative weeks of the largely liquid diet, participants unequivocally expressed that their major challenge stemmed from temptations and cravings for ‘usual foods’. It was evident that the special low-calorie liquid diet was unlike any other diet they attempted in the past and this resulted in varying levels of deviation among participants as they gave into their impulses, particularly in the home eating environment.

> I was really craving food and I found it was harder for me the days when I was home. I am home on my off day and of course you’ve got your little house work and thing to do but you’re not as busy as you would be when you’re at work and stuff. So then it was really, really hard and I had to find things to do to keep my mind occupied; call my husband, call my friends, anybody to talk me out of cheating. [Female participant 1-Focus Group]

As was discovered at baseline, the participants’ temptations were intensified as family members continued with their dietary habits which were seen as ‘triggering’. Some participants’ immediate family members had no regard for the change they were making as no consideration was given to the impact their dietary choices may have had on the participant’s efforts.

> From day 1 it was like if they [the participant’s family members] were conspiring against me. I called that a conspiracy because is like Tuesday before was mackerel, before that was tuna and then all of a sudden in comes out of nowhere the first time in like 6 months they decided ‘oh you start the diet?’ [here’s] fried chicken. [Male participant 1 – Eight week interview]

It was also discovered that the recommended foods for the low-calorie diet proved difficult for some to consume. Many reported that the consumption of non-starchy vegetables became monotonous as they were ill equipped with creative ways for preparing their vegetarian meals. Their creativity with their meal preparation was further stunted due to the availability and accessibility of vegetable options as participants found the cost of vegetables was far too expensive especially at the supermarket.

> So that’s one of the things that had me a bit frustrated cause now I have to buy more vegetables than I normally would have and then seeing how expensive you know it was. I’m like ‘why is it that the things that are good for us so expensive?’ And I mean and I’m complaining right and I am working and my husband is working. How about those who probably have to support a household on their own and have to be on a diet like that you know? Can they afford it you know? [Female participant 1-Focus Group]

Additionally, some participants found it challenging to consume the recommended daily volume of water which led to them experiencing hunger pangs, ultimately resulting in them indulging in foods they were not supposed to consume. Despite these challenges with the recommended diet, most participants preferred the rigidity of this phase of the study as it was a guaranteed way for achieving drastic weight loss-a desired outcome identified by most participants during the baseline interviews. Moreover, participants appreciated the fact that they did not have to “think too much” about the caloric content of the foods consumed. They agreed that the special low-calorie liquid diet made it easier for them to keep calories down and achieve their weight loss.

Despite their initial fears for the low-calorie diet, the participants all agreed that they were particularly concerned about transitioning into the final phase of the study. The flexibility they had with their food choices in the weight maintenance phase, made them quite anxious that they would revert to old habits. Like the other two junctures, food posed the greatest challenge for them during this final phase. Their difficulty in this instance however, revolved around understanding portion and serving sizes, meal planning and staying within the calorie count. To address these challenges, some opted at using calorie counting applications, while others kept food diaries. These two solutions were however short-lived as participants either failed to continuously log foods consumed, or the application they used focused more on westernized foods and very little on Caribbean meals. In the instance of the latter, they used their own estimations to calculate their daily calorie count which may not have been a true reflection of their daily intake.

The cost of the healthy foods continued to be a challenge for some during the six-month weight maintenance phase. Participants struggled with determining which types and combinations of foods they could consume given their financial and caloric constraints. To offset the costs of foods some opted at starting their own kitchen gardens while others received their supply of fresh vegetables from friends and family members who also had kitchen gardens or from vendors who sold them at a cheaper price.

### Situational Barriers

It was generally difficult for the participants to manage their food choices at social events and gatherings. As food plays a substational role in the Barbadian culture, participants noted the heightened peer pressure they experienced while in social settings. From their accounts, it is clear, that culturally they were expected to consume a large volume of food and drink when out at social gatherings. Failure to consume large volumes of food and drink thus resulted in interrogation from their peers as demonstrated in the quote below.

> … the greatest difficulty to manage the diet as such would be only in social settings…You know how Bajans are ‘you aint eating nothing?’ ‘You only eating that?’ So, I guess that’s within this setting, but if you’re outside of this country and you go places people don’t expect you to have a plate full or overflowing of things. [Female participant 2-Eight-week interview]

Furthermore, their failure to consume large volumes of regular food and drink was perceived to be rude and a deviation from the cultural norm.

> …I’ll find other options that will make him [participant’s boyfriend] feel comfortable with me being social in that sense…because he has this thing when we go to people’s houses and I say, ‘thanks no’, ‘you can’t tell people no, they invite you to their house and offer you something you have to take it.’ You know and he says, ‘you can’t go in people’s house and not take what they offer you…they’ll feel that you think what they have is not good enough’. [Female participant 3-Eight- week interview]

In some instances, participants thus felt it best to minimize the number of social events they attended to not appear to be flouting cultural norms. Some were overly critical basing their decision to only attend those events where they felt they had control of decision-making around food consumption. It is interesting to note, though participants were generally tight-lipped about their diabetes diagnosis, some felt it useful to disclose their diagnosis and involvement in the study to address the peer pressure experienced.

The limited healthier food options available at recreational facilities proved problematic for participants even when socializing with persons who were supportive of their lifestyle changes. One participant termed her experience as a “diabetic discrimination” as she recalled the resistance she and her family experienced at the cinema when she attempted to take along her meal replacement shake. It was only when she highlighted her involvement in the study and threats of the ramifications of a drop in her blood sugar levels were made, did they allow it.

There was consensus that all participants experienced shortcomings maintaining their diet because of their daily schedules. It was found that those who had particularly demanding work and family life schedules were more likely to skip meals, purchase unhealthier food on the go and or deal with temptations associated with socializing with their business partners. Furthermore, their high stress work environment often led some participants to emotionally eat (consumed large portions of unhealthy foods). For those participants working in the food industry, the work environment posed a unique challenge as the smell and taste of the foods intensified their cravings. Participants were therefore forced to strategically plan and prepare meals especially during the low-calorie diet phase.

Finally, during the six-month weight maintenance phase, participants noted that they had trouble managing their food choices and addressing peer pressure to indulge in the foods prepared over the Christmas holiday and long weekends with Bank holidays. Culturally, these holidays are celebrated with friends and family with large volumes of food and drink that typically are high in starches and sugars. All participants agreed that their weight and blood sugar levels fluctuated during this phase. This outcome thus propelled some of them to revert to versions of the low-calorie diet.

### Attitudes towards diabetes

At baseline, the participants highlighted their struggles with their diabetes care and management. In addition to their inability to religioulsy take their medication on time, some participants uneqivocally expressed their dislike for taking their medication. For most of them, the fact that they were not expected to take their medication during the course of the study served as motivation to participate and follow the guidelines of the intervention.

It was also observed that participants associated fear with their diabetes. They expressed the heightened sense of anxiety they experience as they think about the complications associated with the illness. One female participant recounted:

> Maintaining this type of eating is really difficult. I am really trying because I have a fear of amputation…I don’t want to lose any of my limbs so I’m trying to stick to a particular way of eating. So I make sure I, you know I stock up on a lot of vegetables and as I said my sister helps me. So she makes sure that I eat what I am supposed to eat. [Female participant 4-Baseline interview]

Despite the fear they associated with the illness, participants usually reverted to their normal eating habits out of frustration generated by not knowing what combination of foods they should consume. When asked how diabetes has affected her daily life, one participant noted:

> It affects [me] due to the things that you see and you hear about the kidney problems and the heart problems and so on. I said ‘it is so dangerous that I wouldn’t like to find that I lose a kidney or so’. So it has got me scared. At first when I heard about it I was like ‘oh shoot I have to come off so much things, what am I going to eat?’ Then when to find out that the majority of the vegetables that you’re eating have sugar, like carrots and so on and you have been eating carrots all the time saying ‘oh it is good for your eyes, oh it’s good’ then to find out that it have a lot of sugar in it. What are diabetes people going to eat when the food have the sugar, the vegetables have the sugar, the fruits have the sugar? What are we going to eat? I does have to question myself ‘look eat something, you have to live too’. So sometimes I fall back on the cake and things like that cause sometimes you don’t know what to eat. [Female participant 5-Baseline interview]

It became more evident that there is a general negative perception of diabetes within the Bardadian culture. During the eight-week diet phase, participants spoke of the sense of embarrassement they felt because of their illness. Though some openly divulged information regarding their illness and by extension the details of their involvement in the study to address peer pressure, there were participants who withheld such information from their co-workers, peers and even family members. Some participants also expressed the reservations their social network members had about them openly sharing information about their illness with others. One male participant during the group interviews noted “I remember people saying ‘But Carlvin you let people know that? You aint shame?’… My mother was actually like ‘but Carlvin I would not let anybody know. You know how Bajans are.”

Furthermore, while rapid weight loss was the main goal of the eight-week diet phase, it became apparent that the public’s negative perception of diabetes influenced the types of comments participants received. Though the quick weight loss was generally perceived as positive for the females the converse was true for the males. During the focus groups the female participants noted that while others commended them for their weight loss, their comments were sometimes flanked by statements as “try and not get too small”. They associated such comments with the culture’s African roots where being voluptuous is perceived as healthy. For the men on the other hand, their rapid weight loss was associated with HIV/AIDS and a loss of their ‘manhood’. The negative comments they received presented them with physchological barriers which they needed to overcome. In some instances, this proved to be a difficult task as it affected how some male participants continued to progress in the study as they were more likely to modify the intervention plan as to not lose too much weight too quickly.

> I think when I first started the study I was either 206 or 207 pounds and now I’m down to 192. So it means that I have lost about 14 or 15 pounds during the last 3 weeks or so. I think if I get down to like say 180, another 12 pounds like that would be my ideal weight if I could keep everything tight and so on. Umm but I really don’t want to go beyond that because then you would excite too much curiosity on the part of the ungodly as to whether or not in your old age you have been with a young girl and captured HIV/AIDS and so on. General foolish comments. [Male participant 2-Baseline interview]

### Social Support

During the baseline interviews all participants agreed that they needed support from their family and peers. They felt that their family members would play a critical role in minimizing the temptations within the household. Although they asserted that they would appreciate constant reinforcement and reminders from their family and peers, there was a thin line between this behaviour being perceived as supportive versus what they deemed as ‘negative support’ (e.g. any support that was given in a chastising manner).

There was a resounding consensus among participants, across the interviews and focus groups that support was an instrumental component for the successful completion of the eight-week diet. Though they differed with respect to their choice of support, it was found that they opted at sourcing assistance from family members, close friends, and co-workers. It was observed that their support systems provided them with assistance in purchasing and or preparing meals, giving words of encouragement, and monitoring their daily food consumption and general well-being. Some participants were fortunate to have their family members and friends join them using a modified version of the low-calorie diet. It is important to note however, others had trouble obtaining support from family members which they believed influenced their abilities to achieve the type of results they wanted. In such instances, they relied heavily on the support of their friends, or their own volition.

Another key source of support for participants were others involved in the study. Many expressed that they drew closer to other participants and members of the research team as they sought emotional, tangible and information support. They all expressed similar opinions on the utility of the weekly visits with the Clinical Investigator and calls by the research assistants. This, they agreed, gave them a sense of accountability, and made them feel as though there were persons who truly cared about their well-being.

Interestingly, as the level of the research team’s support was intentionally changed in the final phase of the study, it was found that participants felt it was easier for them to slip back into their bad habits. When asked how easy or difficult it would be for them to continue with the maintenance phase on their own, one participant stated:

> Difficult in that you know, being that I know I have to come and see Dr. Brown, you want to be on your best behaviour and you do things a certain way. Then you meet the others and you want to be able to say good things too…If that is gone I think it may be a lil easier to slip back into [bad habits] but then you got to remind yourself why you are doing it. It is not for Dr. Brown, it is not for this person, it is for me, it is for myself, it is for my health, you know. And I guess that is where the difficulty [lies]; you would miss that, that part of the support.[Female participant 6-Eight-month interview]

With the absence of the research team’s weekly phone calls and check-ups, the participants created a WhatsApp chat group to motivate one another. Again, this group was deemed an important source of accountability and a forum to share strategies used to overcome any short comings and challenges they experienced during the weight maintenance phase.

Though a change in the support was observed during the final phase of the study, participants’ social network members continued to offer them encouragement, reminders, and joined in their exercise regimes. Participants expressed that the change in support stemmed from the fact the structure of the eight-week low-calorie liquid diet was one where the restrictions and requirements were more quantifiable thereby making it easier for others to provide the types of assistance they required. On the other hand, the scepticism that some faced during their diet was changed into positive feedback and praise. With the participants’ result being more noticeable, it appeared that others were more willing to desist from pressuring them to revert to their bad eating habits while others went as far as to mirror their behaviours.

### Internal drive and self-motivation

The participants’ willpower proved particularly useful during the eight-week very low-calorie diet. They all acknowledged that though social support was necessary at times, their ability to successfully complete the low-calorie liquid diet was solely up to them. Their level of self-motivation grew as they saw their weekly results (weight loss and reductions in blood sugar and blood pressure levels). Some were particularly motivated by the fact that they were no longer dependent on medication to manage their different chronic illnesses. More importantly, most participants’ self-efficacy improved by the end of the eight-week period as they believed the rigidity of the study’s restrictive diet helped them to develop new eating habits needed for the considerable weight loss they failed to achieve in the past. Although no gender specific differences were found, it was unearthed that participants who incorporated exercise during the low-calorie diet, researched foods and suitable portion sizes were better equipped to address the challenges of the diet. These participants felt better overall (mentally and physically), and they were generally the ones who saw the biggest changes in their baseline measurements.

It is also worthy to note that there was a concomitant improvement in most participants’ ability to resist temptations as their perceived self-efficacy grew. Participants planned, prepared, and pre-packaged their meals to take with them when they were on the go; some even utilized this strategy when attending social events. Most reported that they did not previously pre plan nor pay attention to the types of foods consumed. As such, this strategy proved quite useful as it helped to ensure that participants stuck to their restrictive diet.

Participants who did particularly well in maintaining their weight, blood glucose, and blood pressure levels highlighted that this success was due to their consistent research on the optimal combination of food groups, portion sizes and daily caloric intake. They consciously made efforts to increase and maintain their consumption of vegetables, measure their foods using either a kitchen scale and or measuring cups, examine the nutritional information on food labels and calculate the number of calories expended daily.

Given the greater freedom in their food consumption choices, many spoke of their desire to maintain the practices and behaviours developed over the first two months of the study to “get rid” of their diabetes. Their strong dislike for taking medication and the results they obtained served as a catalyst for their drive to maintain a healthy lifestyle. Moreover, some participants spoke of their desire to serve as an example for others in their family and the wider society at large. In the instance of the former, participants saw the need to demonstrate to others in their inner circles that they could successfully complete the study and reap the benefits of their efforts. With the high prevalence of diabetes in Barbados, they felt that by sticking to the programme they would be better equipped to assist other citizens living with the disease.

## Discussion

This study provides insight into the challenges and facilitators experienced by 25 Barbadians who participated in an intervention aimed at type 2 diabetes remission. While the current study shares similarities with the findings of Rehackova et al.[20], our analysis revealed more culturally related situational barriers across various settings and a negative attitude towards type 2 diabetes as predominant challenges. The participants’ internal drive and self-motivation along with social support facilitated their adherence to the low-calorie liquid diet and structured weight maintenance over a six-month period. Despite the excessive use of self-control rhetoric in the literature regarding diabetes management, our data supports Albert Bandura’s assertion that most human behaviour is determined by multiple interacting factors [26]. The data clearly demonstrates the interconnectedness of intrapersonal and interpersonal influences for type 2 diabetes remission within the Barbadian context.

## Findings in context

Based on the Social Ecological model, behaviour change is maximised when there is an interaction between the multiple levels (intrapersonal, interpersonal, organizational, community) of influence on specific health behaviours [27]. Our findings elucidate the wider cultural norms with respect to food consumption within the Barbadian social setting. The participants’ wishes to refrain from certain foods and drink were usually met with resistance by others around them, as food is a central aspect of the culture. Moreover, food has significant symbolic and social meanings as it revolves around the bonds created and nourished through the preparation and consumption of meals [28-31]. In this regard, it plays an integral role in the maintenance of familial and social networks. Participants were thus faced with heightened levels of temptation, especially during the formative weeks of the intervention, to consume ‘regular foods’ as culturally they were expected to consume large volumes of food and drink when out at social gatherings. Their levels of perceived control in these settings were constantly challenged as they progressed through the phases of the study. In some instances, they experienced great difficulty in maintaining control, giving into the peer pressure and as such negatively impacting the behaviour required to reverse their diabetes.

Furthermore, the findings of this study mirror other qualitative research which suggest that social functions impede the dietary alterations required for effective diabetes care [13, 32-36]. The participants’ refusal to consume large volumes of regular food and drink was not socially acceptable and was considered to be a deviation from cultural norms. Peer pressure worsened if they did not wish to disclose their diabetes as their reason for what seemed to others to be unusual consumption behaviour. Though some were generally tight-lipped about their diabetes diagnosis, some felt it useful to disclose their diagnosis to address the peer pressure experienced. Members of their social network however, appeared to be more willing to accept their wishes to refrain from ‘regular foods’ only after their involvement in the study was disclosed. Nonetheless, the data is indicative of the high level of resiliency and the positive temperament towards problem solving and logical decision making required to successfully manage the behaviour required for type 2 diabetes remission.

Family dietary behaviours also represents one of the major challenges for persons with type 2 diabetes [16, 37-39]. Within the home environment, the data revealed the heightened levels of temptations participants faced on a regular basis and the overt ridicule and sabotage they experienced as they did their best to engage in the recommended behaviour. Such stigma and ridicule thus increase the likelihood that the requisite self-care behaviours for remission may be compromised [40, 41]. Furthermore, consistent with previous research, the challenges participants experienced making the requisite dietary changes were intensified if their family members were not necessarily willing to follow their diet regimen and at the congregate meal sites they continued eating their regular meals [16, 35, 39, 42, 43]. Indeed, the integration of their new dietary habits with their families culturally established dietary habits and preferences presented a major barrier to their dietary change and adherence [16, 44, 45]. Diabetes-friendly substitutes and cooking methods appeared less appealing and compounded the participants’ temptation when we consider their traditional cooking methods of using saturated fats for frying and sugar for stewing meats for example. Having difficulty with finding appealing ways to prepare their meals, participants therefore complained of the monotony of the foods they ate during the low-calorie diet; some resorted to simply eating their vegetables raw while others ate them in soups. Again, in these instances, the urge to consume regular foods was further intensified and lead to lapses in their dietary adherence.

The participants’ narratives revealed that while modifying their dietary behaviour, female participants were faced with a unique challenge as they were still responsible for purchasing and preparing meals for their family. In this regard, their needs were secondary to those of the other members of the household as they had a sense of obligation towards fulfilling their dietary wishes and needs. While the male participants may have had their meals prepared by their spouses or significant others, there was limited or little control over foods prepared. Although this finding maybe suggestive of a gendered problem, more research is required to confirm the impact of the family’s influence on the diet modification efforts Barbadian males and females are required to make to reverse their diabetes.

In the instance of the work environment, some faced challenges balancing their erratic work schedules with the demands of their family life and that of the study. Their irregular work hours primarily adversely impacted their ability to eat on time. In some instances, participants either missed meals or they were forced to consume unhealthy foods as these were easier and more convenient to grab on the go. Social events and meetings within the work setting also proved particularly problematic as more often than not unhealthy foods inclusive of fried foods, cakes and pastries were at the crux of these gatherings. To minimize social isolation and the negative impacts associated with a decline of foods offered, they sabotaged their progress by partaking in the foods served. This finding thus suggests that the participants’ colleagues were not necessarily sensitive to their needs and the work environment presents significant barriers which can have deleterious effects on their diabetes remission efforts.

The limited healthy food options provided at recreational facilities also demonstrate a greater need for facilitative environmental changes that can drive the healthy lifestyle behaviours required to effectively manage and or reverse one’s diabetes. With an overabundance of low-cost, high energy dense manufactured foods and an adoption of a Western oriented diet, it is not surprising that there is an upward shift towards the production of foods that are high in fats and calories and low in fibre, vitamins, and minerals [46, 47]. The country’s import policies and high food importation bills are indicative of its colonial legacy and intrigue for imported foods which are perceived to be of a superior quality, cheaper, and trendier than locally produced foods and a symbol of one’s social status [48, 49]. Consequently, this resulted in the morphing of an environment characterized by a proliferation of cheap foods with low nutritional value, while more appropriate, healthier options for those with diabetes are high in cost [24]. If participants are to maintain their newly adopted healthier lifestyle, the wider environment must support healthful choices [50-54]. As asserted by the aforementioned authors, the skills garnered over the course of this intervention would be of little utility if individuals do not have access to healthy foods and safe settings for physical activity. The absence of such resources thus impedes one’s ability to maintain the healthy diet and healthy levels of physical activity required for type 2 diabetes remission.

### New perspectives and future interventions

The significance of food in the Barbadian culture represents a major consideration in the remission of type 2 diabetes. Future interventions need to provide participants with information and strategies which are specific to their preferences and needs. More specifically, participants need to be taught how to incorporate traditional foods and cooking practices in their new dietary regime, how to read and make sense of food labels, how to prepare healthy meals when they have limited time and or cooking skills and how to address social gatherings, cravings and slip ups they may experience [44, 45, 55, 56]. These strategies will thus aid in minimizing the stressors associated with them deviating from the cultural norms and social isolation they may experience as a result of not being fully able to participate in social occasions because of their dietary needs.

It is understandable that participants are faced with multiple barriers due to the diverse activities they engage in daily. Rooted in an ecological perspective, it is crucial that the routines, resources, and activities of the participants’ households are considered and accommodated as they make efforts towards behaviour change [45, 57, 58]. To this end, it would prove immensely useful to intimately involve members of the participants’ social network (particularly their family and friends) as they have crucial roles in their decisions. From an intrapersonal level, it is critical that individuals are taught concrete behaviours and skills with an aim towards empowering and providing them with autonomy in the decision-making processes associated with the health behaviours required for type 2 diabetes remission.

There is a need for ongoing follow-up and support for behaviour change. As was seen with the study’s weekly visits, the Clinical Investigator provided critical assistance in refining the participants’ problem-solving skills, she encouraged them when their test results were less than perfect and guided them on how to respond to new issues that emerged. It was not surprising that participants felt a high degree of accountability to the chief investigator. Their concerns about their ability to effectively transition to the maintenance phase is indicative of their desire for continuous reinforcement and formal support which they perceived to be crucial for their successful completion of the study. In the absence of the formal support provided by the research team, they relied on one another. When they deviated off course, they drew on encouragement from each other; some modelling their behaviours to that of other participants who successfully devised strategies for addressing challenges faced. This finding thus demonstrates the utility of having others around who can demonstrate the small steps that can be taken to achieve a complex goal.

The data underscores the importance of an interpersonal level of influence to aid in the improvement of the participants’ self-efficacy. Though they differed with respect to their choice of support it was found that participants opted at sourcing assistance from family members, close friends, members of the research team inclusive of other participants and co-workers. Most however stated that family members were their major support system, providing them with assistance in purchasing and preparing meals, giving words of encouragement, and monitoring their daily food consumption and general wellbeing. This finding echoes the work of Vassilev et al. [59] which illustrates the critical role of family members in the management of one’s chronic illness. However unlike Vaccaro et al. [60] and Barrera et al. [61] who posited that there was a strong correlation between family social support and diabetes self-management among ethnic minorities, the preliminary data does not coincide with these results. Though participants of the study are generally members of a homogenous ethnic group, some had trouble obtaining consistent supportive behaviour from family members whom they believed influenced their abilities to achieve the type of results they wanted.

Whilst the abovementioned level of influence proved useful in assisting participants to overcome challenges faced, the intrapersonal level was perceived to be a critical facilitator throughout the lifespan of this intervention. This level of influence was indicative of the grave responsibility participants had, not only to induce lifestyle changes by joining the study, but also to maintain the healthy alternatives they adopted as they progressed through the intervention. Though difficult to achieve in practice, they took charge of developing strategies that aided in the improvement of their self-efficacy. Consequently, through strategic goal setting, planning and pre-preparation of meals, participants overcame daily challenges. More importantly, by doing such they were able to meet their goals which was evident in the sustained reduction of their baseline measurements.

### Strengths and limitations

Being the first study of its kind to be conducted in a Global South setting among an Afro-Caribbean population, this study provides unique insights into the barriers and facilitators to type 2 diabetes remission. It highlights the reciprocal interactions between intrapersonal, social, and environmental variables and the deep-seated impact of cultural influences on one’s efforts for dietary induced diabetes remission. Although the experiences of a small sample of participants were captured, this study provides valuable information for the development of future diabetes remission interventions in Barbados and has implications for similar programmes in other countries of the Caribbean. However, given the study’s rigidity (e.g., provision of meal replacements, structured monitoring, and provision of support from the intervention team), it is unclear if similar results would be obtained in primary healthcare settings where similar resources may not necessarily be available.

## Conclusion

This study examined the barriers and facilitators 25 Barbadians experienced as they progressed through three phases of a diabetes reversal feasibility study. The data revealed that there were interrelated and overlapping barriers including the social significance of food and the associated temptations and family and social support. In the same vein, social support received from participants’ social relationships and members of the study was deemed critical for their successful completion of the intervention. At the intrapersonal level of influence, the participants’ self-efficacy and determination was an equally important facilitator for the completion of the intervention.

## Data Availability

The data that support the findings of this study are available on request from the corresponding author (NU).

## Acknowledgements

We wish to acknowledge the contributions of the Barbados Diabetes Association, the Barbados Diabetes Foundation, and the George Alleyne Chronic Disease Research Centre. For their assistance with the data collection, we thank Dr. Karen Bynoe, Andre Greenidge, Melissa Abed, and Krystal Boyea. Finally, we thank Professor Roy Taylor, the study’s Co-principal Investigator, who assisted in establishing the study and the protocols for the very low-calorie diet phase and the metabolic outcome measures.

